# Role of monocytes in endothelial glycocalyx shedding during Puumala orthohantavirus infection

**DOI:** 10.1101/2024.08.25.24312546

**Authors:** Luz E. Cabrera, Isaac Polanco, Johanna Tietäväinen, Satu Mäkelä, Olli Vapalahti, Antti Vaheri, Jukka Mustonen, Tomas Strandin

## Abstract

Vascular leakage characterizes Puumala orthohantavirus (PUUV)-caused hemorrhagic fever with renal syndrome (HFRS). Disruption in the endothelial glycocalyx layer, which protects blood vessels from increased vascular leakage into tissues, may contribute to disease severity during acute PUUV-HFRS. Recent evidence suggests that increased heparanase (HPSE) activity could play a role in glycocalyx degradation. Additionally, dynamic changes in monocyte populations, especially a decrease in endothelium “patrolling” CD14^+^CD16^+^ nonclassical monocytes (NCMs) are observed in acute PUUV-HFRS. To investigate HPSE expression levels in different monocyte subsets and their relationship to the numbers of circulating endothelial cells (CECs) as marker of glycocalyx degradation during PUUV-HFRS, we analyzed patient peripheral blood mononuclear cells collected at acute and recovery stages of the disease by flow cytometry. CECs were significantly increased in acute PUUV-HFRS patients, and gradually decreased towards the postacute and recovery phases. Moreover, we identified significant correlations between CECs, the frequencies of different monocyte subsets and their HPSE expression, and clinical parameters. The decrease in HPSE expressing NCMs correlated with plasma HPSE levels, indicating a potential role for monocytes in modulating endothelial glycocalyx shedding. Furthermore, co-culture experiments suggested PUUV-infected endothelial cells (ECs) as regulators of monocyte HPSE expression. The study enhances understanding of EC and monocyte dynamics during PUUV-HFRS, providing insights into immune responses and potential vascular complications. Subtle variations in monocyte subsets emphasize their possible variable roles in disease progression and changes throughout the disease course. Overall, the study contributes to unraveling the complex interactions between viral infections, immune responses, and vascular dynamics, guiding future investigations and interventions in PUUV-HFRS-associated vascular complications and glycocalyx shedding in the kidneys.

## Introduction

The dynamic interplay between viral infections and the vascular system has been a subject of increasing interest, particularly in the context of viral diseases with diverse clinical manifestations (Fosse et al., 2021). Among these we find Puumala virus (PUUV), which causes a relatively mild form of hemorrhagic fever with renal syndrome (HFRS) (Vaheri et al., 2023). Despite the non-cytopathic effect of PUUV infection in endothelial cells (EC) and monocytes (Temonen et al., 1993), which play a key role in vascular integrity, the infection leads to remarkable vascular complications evidenced by capillary permeability, increased coagulation and fibrinolysis, thrombocytopenia and shedding of the endothelial glycocalyx (EGC) components (Hepojoki et al., 2014).

The EGC disruption is proposed as a potential factor driving capillary leakage in acute HFRS (Connolly-Andersen et al., 2014; Tietäväinen et al., 2021; Du et al., 2023). The EGC acts as a crucial interface between circulating blood components and the endothelium (Reitsma et al., 2007), maintaining the vascular integrity and influencing its permeability. The enzyme heparanase (HPSE) cleaves heparan sulfate molecules and acts as a major mediator in the degradation of the EGC (Becker et al., 2015; Masola et al., 2022). The role of HPSE in compromising vascular integrity in viral diseases has been recognized, including PUUV-HFRS (Kinaneh et al., 2021; Cabrera et al., 2022; Drost et al., 2022). Further, EGC damage has been proposed as a mechanism linking hyperglycemia with disease severity in HFRS (Tietäväinen et al., 2021). During other viral diseases such as COVID-19, altered vascular permeability may be responsible in the detachment of ECs associated with disease severity (Guervilly et al., 2020; Varga et al., 2020; Kinaneh et al., 2021; Drost et al., 2022). This detachment not only facilitates the dissemination of the virus but also impairs vascular reactivity and nitric oxide (NO) production, crucial for blood vessel relaxation and vascular homeostasis by preventing coagulation and inflammation.

Monocytes, as key players in the immune response, exhibit remarkable heterogeneity, with distinct subsets playing different roles in immune response modulation, tissue homeostasis, and pathogen defense (Williams et al., 2023). Notably, distinct monocyte subsets assume varying roles in both steady-state and pathological conditions. Classical monocytes (CM, expressing CD14^++^CD16^-^) primarily initiate innate immune responses, engaging in phagocytosis and migration through the expression of chemokines, scavenger receptors, and proinflammatory cytokines (Gren et al., 2015). Intermediate monocytes (IM, CD14^++^CD16^+^), on the other hand, are linked to antigen processing, presentation, monocyte activation, inflammation, and differentiation (Zawada et al., 2011), and nonclassical monocytes (NCM, CD14^+^CD16^+^), are unique in their patrolling behavior, particularly in surveilling vasculature and thus interacting with the endothelium (Cros et al., 2010). Importantly, monocyte translocation through vascular bed requires EGC degradation mediated by HPSE, which is expressed on monocyte cell surface (Sasaki et al., 2004) and suggests that monocytes are major source of HPSE activity.

In the context of acute PUUV-HFRS, our previous research revealed dynamic changes in monocyte populations. NCMs are significantly diminished in the circulation during acute PUUV-HFRS by rapidly redistributing to affected tissues during infection, accompanied by an increase in CMs and IMs in blood (Vangeti et al., 2021). This NCM redistribution leads to inflammation and tissue damage, particularly in the kidneys during acute PUUV-HFRS, suggesting their involvement in renal pathology.

In the present study, we hypothesized that monocyte populations may play a crucial role in the increased expression of HPSE, thereby contributing to the breakdown of the endothelial glycocalyx and exacerbating vascular complications in PUUV-HFRS (Sironen et al., 2017). By investigating the intricate mechanisms involving monocyte subsets and HPSE expression, our research aims to shed light on the complex interplay between immune responses and vascular complications during PUUV infection. These findings not only contribute to a deeper understanding of the pathophysiology of this disease, but also lay the foundation for potential therapeutic interventions in the future, to alleviate the vascular complications associated with this viral illness.

## Materials & Methods

### Ethics statement

The Ethics Committees of Tampere University Hospital (permit number R04180) and Hospital District of Helsinki and Uusimaa 100 (HUS/853/2020, HUS/1238/2020) approved the use of patient samples. All subjects gave written informed consent in accordance with the Declaration of Helsinki.

### Patient population

The study material consisted of plasma, urine, and PBMCs from hospitalized, serologically confirmed acute PUUV infection at Tampere University Hospital, Finland, between 2005 and 2009. Patient samples were collected from a cohort of 25 individuals at different time points, which included the acute phase of the disease for samples taken during hospitalization, the postacute/convalescent phase for samples taken 15 days after discharge, and the full recovery phase for samples taken 6 months and one year after the acute phase. Full recovery phase samples were used as controls. Standard methods were employed to determine daily white blood cell (WBC) count, plasma C-reactive protein (CRP), and serum creatinine concentrations at the Laboratory Centre of the Pirkanmaa Hospital District (Tampere, Finland). Details about the clinical and laboratory data of the patient cohort can be found in Table 1.

**Table 1.**
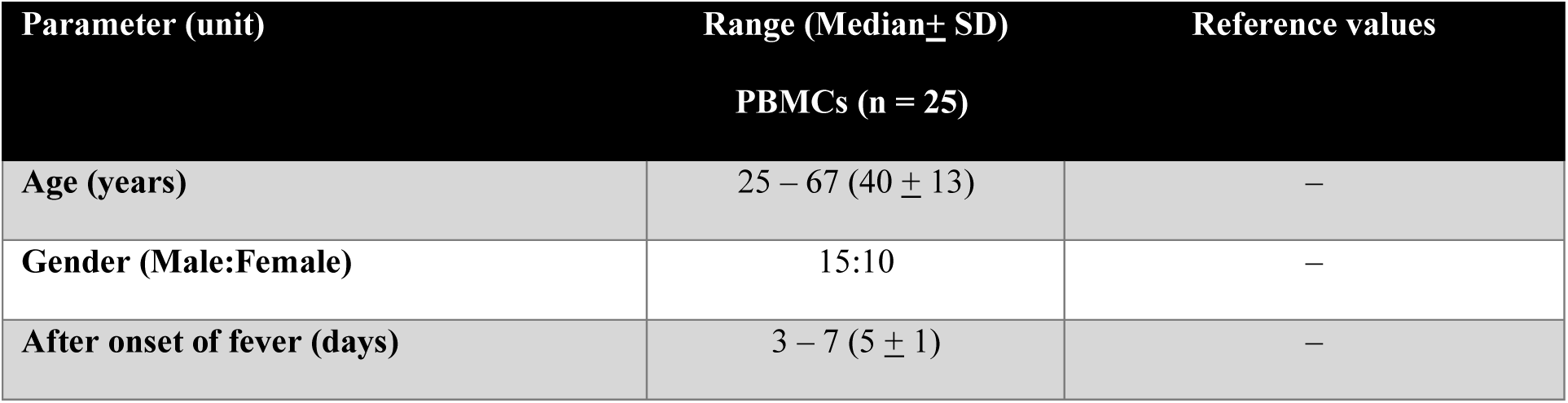
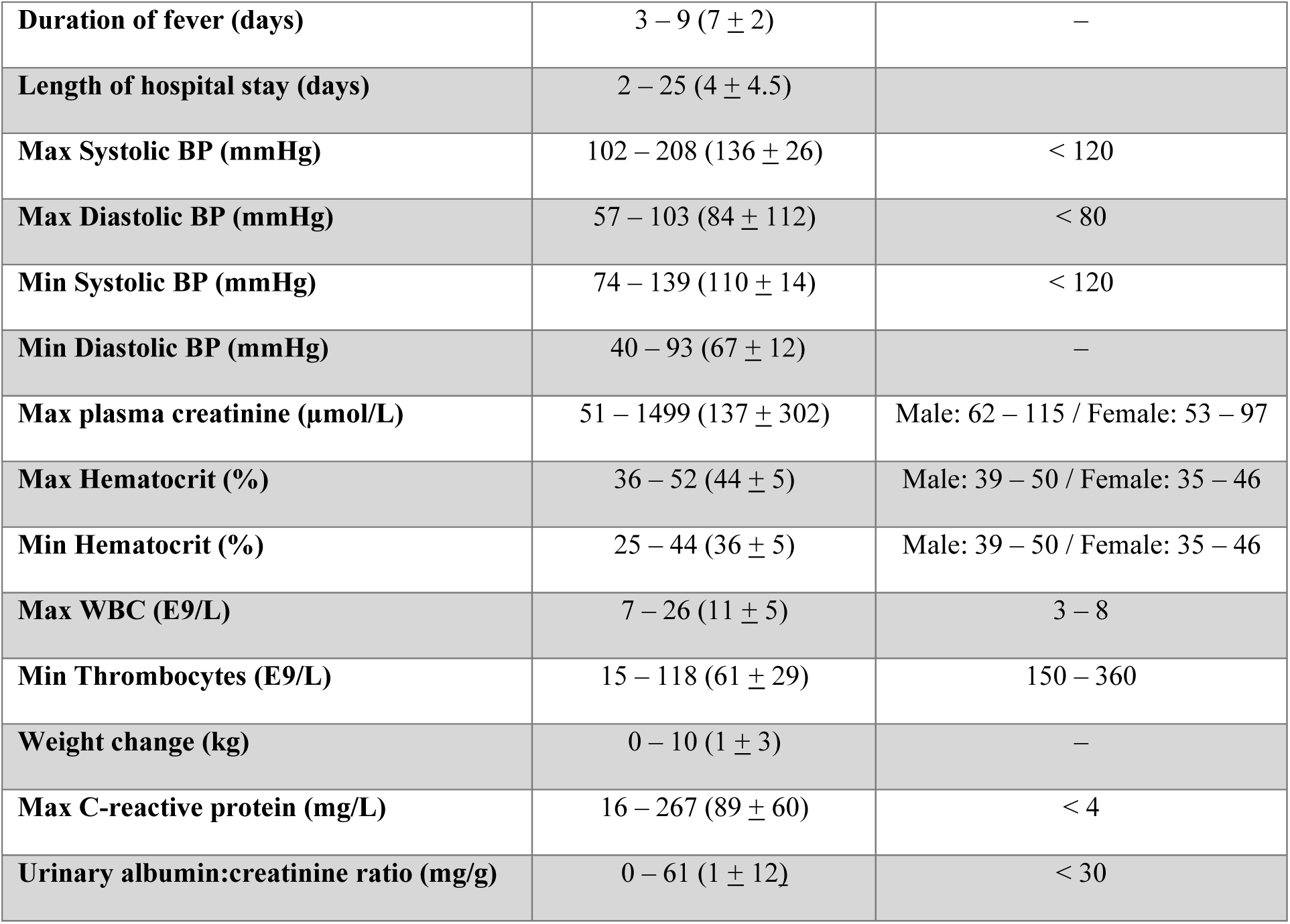
Clinical and laboratory data during hospitalization in patients with acute PUUV infection. BP = blood pressure; WBC = white blood cells; max = maximum; min = minimum.

### Flow cytometry

Frozen PUUV patient PBMCs were thawed in a 37°C water bath for 5 min, then washed with RPMI-1640 (Sigma Aldrich) supplemented with 10% inactivated FCS, 100 IU/ml of penicillin, 100 μg/ml of streptomycin, 2mM of L-glutamine, and 100 μg/ml DNAse I (Sigma Aldrich). After a 10 min incubation at RT, the cells were washed once with the conditioned medium and once with PBS-EDTA (PBS with 2 mM EDTA), followed by cell quantification using Bio-Rad cell counter TC20.

One to three million PBMCs were incubated in 1% FCS and FcR blocking reagent (BioLegend), and the cells stained for 30 min at RT with a cocktail of fluorescent-dye conjugated anti-human mouse mAbs recognizing cell surface antigens and a dead cell marker (antibody panel detailed in Table 2). After staining, the cells were washed with PBS-EDTA and fixed with 1% paraformaldehyde before FACS analysis with a 4-laser (405, 488, 561 and 637 nm) Quanteon Novocyte (Agilent Technologies). The FACS data was analyzed with FlowJo 10.8. The datasets were normalized independently and concatenated. Cell populations were identified by Uniform Manifold Approximation and Projection for Dimension Reduction (UMAP), allowing for their unsupervised identification, based on the detection of different fluorochromes used in the flow cytometry panel. To generate UMAP plots, the minimum distance was set at 0.5 and the nearest neighbors’ distance was set at 15, using a Euclidean vector space. Clusters were then identified using the phenograph plugin.

**Table 2.** Flow cytometry antibody panel.

| Marker | Fluorochrome | Clone | Company |
| --- | --- | --- | --- |
| CD123 | PE | 6H6 | BioLegend |
| CD19 | PE | LT19 | Immunotools |
| CD3 | PE | OKT3 | BD Biosciences |
| CD56 | PE | B159 | BD Biosciences |
| Heparanase 1 | FITC | HPA1 (E-10) | Santa Cruz Technology |
| Live/Dead | FVS 570 | – | BD Biosciences |
| CD14 | PE-Cy7 | M5E2 | BD Biosciences |
| CD34 | APC | 4H11 | Invitrogen |
| CD45 | V450 | 2D1 | Invitrogen |
| CD133 | Brilliant Violet 605 | TMP4 | Invitrogen |
| CD146 | Brilliant Violet 711 | P1H12 | BioLegend |
| CD16 | Brilliant Violet 786 | 3G8 | BD Biosciences |
| HLA DR | BB700 | G46-6 | BD Biosciences |
| CD138 | APC-CY7 | 44F9 | MACS Miltenyi |

### *In vitro* coculture

Blood microvascular endothelial cells (BECs) were obtained from Lonza and maintained in endothelial basal medium (EBM-2) supplemented with SingleQuots™ Kit containing 5% fetal bovine serum (FBS), human endothelial growth factor, hydrocortisone, vascular endothelial growth factor, human fibroblast growth factor-basic, ascorbic acid, R^3^-insulin like growth factor-1, gentamicin and amphotericin-B (Lonza). For experiments the cells were used at passage 8. BECs were infected for 3 days with live or UV-inactivated PUUV (strain Suo). Multiplicity of infection was 1 as calculated by using Vero E6 cells. Monocytes were isolated from PBMCs obtained from healthy volunteers using CD14 magnetic beads (Miltenyi Biotec) according to the protocol provided by the manufacturer. Isolated monocytes were co-cultured with infected BECs in a 3:1 ratio (with monocyte concentration of 1 million/ml) for 24-hr. Cell culture supernatant was collected after which cells were pelleted by centrifugation 400g for 5 min and stained for flow cytometry as described above.

### ELISA

Syndecan-1 was measured from urine of patients using an ELISA kit purchased from R&D systems following the manufacturer’s protocol (Human Syndecan-1 DuoSet, Catalog #: DY2780; Bio-techne, Abingdon, UK).

### HPSE Measurement Assay

HPSE levels from patient plasma and urine was measured through the enzyme’s ability to cleave HS, which was quantified with the use of a standard curve. In detail, Nunc maxisorp flat-bottom 96-well plates (Thermo scientific; Breda, The Netherlands) were coated with 10 μg/mL heparan sulfate from bovine kidney (HSBK) (Sigma– Aldrich; Zwijndrecht, The Netherlands) in coating buffer (3.3 M ammonium sulfate ((NH_4_)_2_SO_4_)), for 1h at 37 °C. Subsequently, plates were washed with PBS supplemented with 0.05% Tween 20 (PBST) and blocked with 1% BSA in PBS at RT. After blocking, plates were washed with PBST, followed by a final washing step with PBS. Plasma and urine samples were then incubated for 2 h at 37 °C, in a 1:4 dilution in HPSE buffer (50 mM citric acid–sodium citrate, 50 mM NaCl, 1 mM CaCl2 at pH 5.0). Next, plates were washed with PBST and incubated with primary mouse anti–rat IgM HS antibody JM403 (Amsbio; Abingdon, United Kingdom, cat. no. #370730–S, RRID: AB_10890960, 1 μg/mL in PBST) at RT for 1 h, washed with PBST and then incubated with secondary goat anti–mouse IgM HRP antibody (Southern Biotech; Uden, The Netherlands, cat. no. #1020–05, RRID: AB_2794201, 1:10,000 dilution in PBST) for 1h at RT and washed with PBST once again. Finally, 3,3′,5,5′–tetramethylbenzidine (TMB) substrate (Sigma–Aldrich, Zwijndrecht, The Netherlands) was added to the plates, the reaction was stopped by the addition of 0.5 M sulfuric acid, and absorbance was measured at 450 nm. The HPSE concentration in the plasma and urine of patient samples, as well as from podocyte cultures, was compared to a standard curve of recombinant human HPSE (Bio-techne; Abingdon, UK, Cat#7570–GH–005). Urine concentrations measured were then normalized by dividing the values by creatinine (heparanase:creatinine ratio).

### Statistical analyses

Statistical analyses were performed using GraphPad Prism 8.3 software (GraphPad Software, San Diego, CA, USA). Statistically significant correlations between parameters were assessed by calculating Spearman’s correlation coefficients, and differences between groups were assessed with Kruskall-Wallis or ordinary 2-way ANOVA analyses, depending on sample distribution and the number of groups analyzed.

## Results

### Circulating mature and progenitor endothelial cells are increased during acute PUUV-HFRS

Quantitative analysis of peripheral blood mononuclear cells (PBMCs) using flow cytometry (gating strategy shown in Fig. 1A) revealed dynamic alterations in endothelial cell (EC) counts across different stages of PUUV-HFRS. During the acute phase of PUUV-HFRS (sampling at 1^st^ day of hospitalization), a statistically significant increase in EC counts (both circulating endothelial cells, CECs, and circulating endothelial progenitors, CEPs, Fig. 1B-C) was observed compared to healthy recovery stage (12 months post-infection). The EC counts then gradually decreased in the postacute phase but remained slightly elevated compared to disease resolution. Notably, in the recovery stages at 6 months and 12 months post-infection, EC counts reached their lowest levels, with no significant difference between these two time points. These data suggest compromised endothelial integrity in PUUV-HFRS.

**Figure 1.**
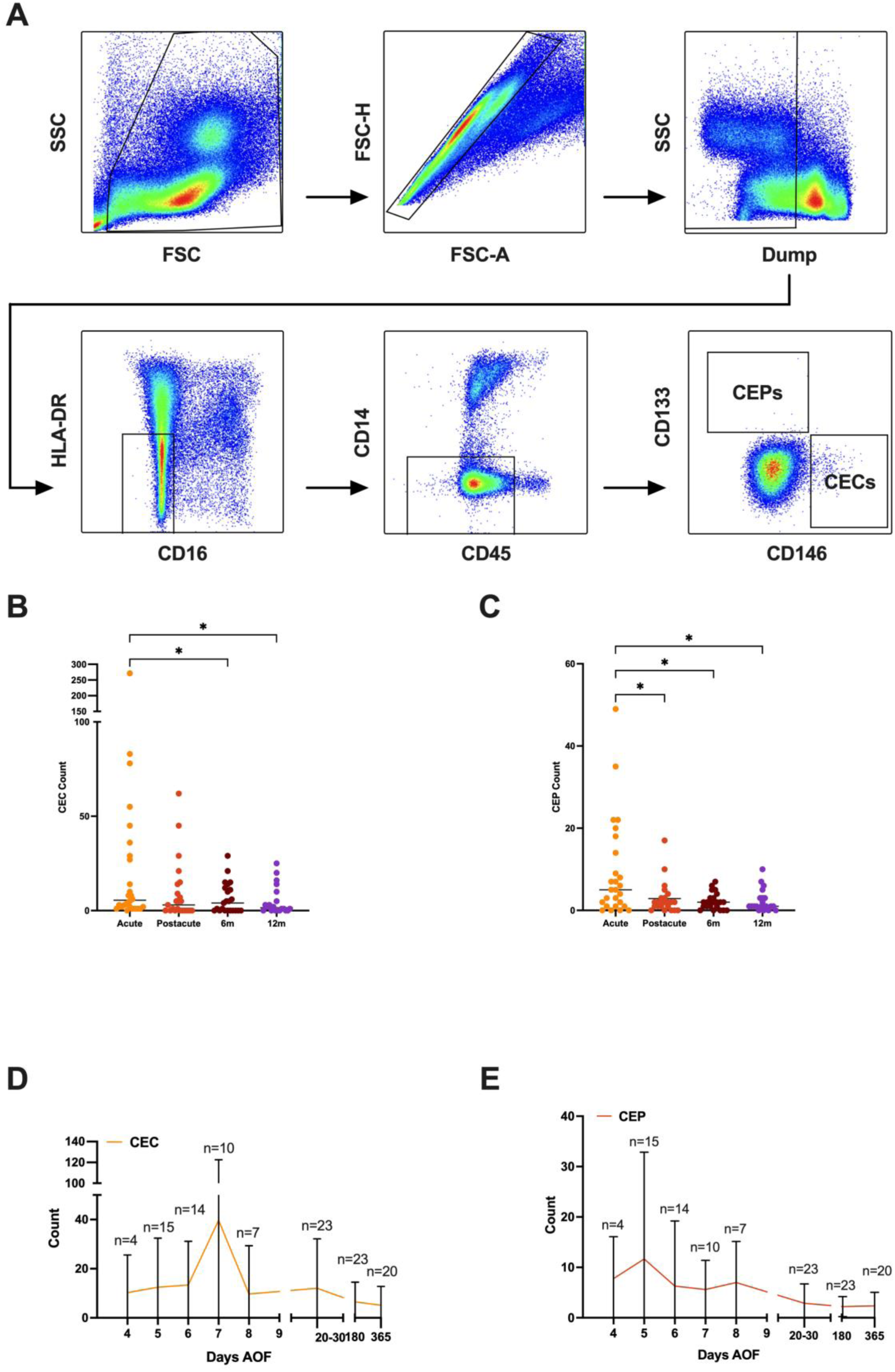
Identification and quantification of endothelial cells in acute PUUV-HFRS Patients. (**A**) Flow cytometry gating strategy for the identification of endothelial cells (ECs) from peripheral blood mononuclear cells (PBMCs). The gating strategy involves a stepwise selection process, starting with morphological gating for debris exclusion (FSC vs. SSC plot), followed by singlet gating to exclude doublets (FSC-A vs FSC-H), and a dump channel for the exclusion of dead cells and CD3, CD56, CD19 expressing cell populations. Subsequently, a double-negative gate for HLA-DR and CD16 is applied, followed by a CD14^-^/CD45^-/dim^, to ensure the exclusion of background when identifying rare cells such as ECs. Then, the final gate plots CD133 and CD146 markers, to identify the circulating EC populations of interest: CD133^-^/CD146^+^ for mature CECs and CD133^+^/CD146^-^ for CEPs. (**B-C**) Graph depicting individual counts of (**B**) Mature CD146^+^/CD133^-^ CECs and (**C**) CD146^-^/CD133^+^ CEPs obtained from flow cytometry analysis. The x-axis illustrates different groups: acute, postacute, and recovered (6 months and 12 months post-infection) PUUV-HFRS patients. Each data point represents a CEC count from one individual within the specified group. (**D-E**) Timeline graphs of the (**D**) CEC and (**E**) CEP cell count means (± SD) from PUUV-infected patients during acute, postacute, and recovery phases. *p < 0.05; p values calculated with Kruskall-Wallis test. *EC = endothelial cell; CEC = circulating endothelial cells; CEP = circulating endothelial progenitors; AOF = after onset of fever; FSC = forward scatter; SSC = side scatter; PUUV = Puumala orthohantavirus; PUUV-HFRS = Puumala orthohantavirus-caused hemorrhagic fever with renal syndrome; SD = standard deviation*.

The acute PUUV-HFRS patient samples were further stratified based on days after onset of fever (aof) at the time when samples were collected. This allowed us to observe kinetic differences between CEC and CEP responses (Fig. 1D-E). Surprisingly, the peak increase in CEPs occurred earlier as compared to CECs (5 vs. 7 days aof), which suggests either that vascular integrity is being compromised before detectable CEC response occurs in our assay or that CEPs mature directly into CECs during acute PUUV-HFRS.

### Dynamic modulation of monocyte subpopulations and HPSE expression during PUUV infection

Following the identification of monocytes using flow cytometry gating (Supplementary Figure 1), these immune cells were gated further into their well-known subpopulations of classical, intermediate, and nonclassical monocytes (CM, IM and NCM, respectively), based on their CD14 and CD16 expression (Figure 2A). Moreover, the assessment of their frequency changes across different stages of PUUV-HFRS revealed a significant decrease in the frequency of NCM during acute PUUV-HFRS, similarly as previously published (Vangeti et al., 2021), compared to all the other groups (Figure 2A-B). Meanwhile, no visible distinctions are observed in IM and CM frequencies between the PUUV-HFRS groups, with CMs showing consistently high frequencies in all groups.

**Figure 2.**
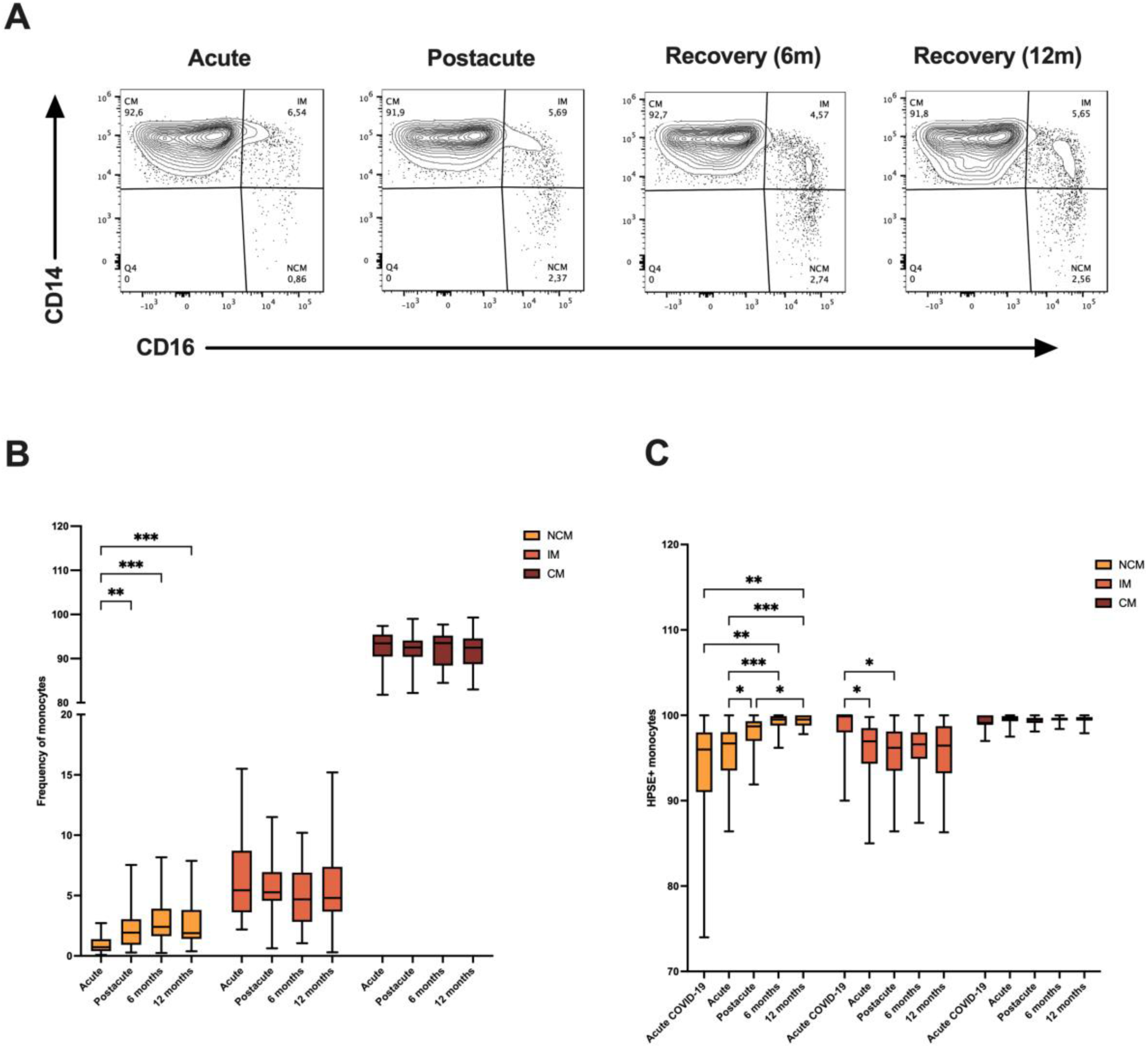
Monocyte subpopulation frequencies and HPSE expression in different stages of PUUV-HFRS. (**A**) Contour plots of all monocytes from concatenated PUUV-HFRS samples, depicting the distribution of classical, intermediate and nonclassical monocyte subpopulations in acute, postacute, recovery (6- and 12-months post-infection) stages of PUUV-HFRS. The quadrant gate depicts the variable frequencies of CM, IM, and NCM in each disease stage. (**B**) Box plots illustrating the frequency distribution of CM, IM, and NCM across different disease stages (acute, postacute, recovery including 6- and 12-months post-infection). The y-axis represents the percentage of monocytes, with each subpopulation displayed in distinct colors. (**C**) Box plots showing the expression levels of the HPSE surface marker in monocyte subpopulations across the different disease stages. *p < 0.05, **p < 0.01, ***p < 0.001, ****p < 0.0001. P values calculated with 2-way ANOVA Tukey’s multiple comparisons test. *PUUV-HFRS = Puumala orthohantavirus-caused hemorrhagic fever with renal syndrome; HPSE = heparanase; CM = classical monocyte; IM = intermediate monocyte; NCM = nonclassical monocyte*.

Additionally, the analysis of HPSE expression in monocyte subpopulations displayed distinct patterns. Samples from acute PUUV infection exhibited significantly lower HPSE expression in NCM, compared to postacute and recovery stages of the disease. However, no significant differences are observed between PUUV-HFRS stages in IM. For CM, HPSE expression remained consistently high across all disease stages, with no significant differences observed.

These findings suggest a dynamic modulation of monocyte subpopulations and HPSE expression during viral infections. The pronounced decrease in NCM frequency and HPSE expression in acute PUUV-HFRS compared to postacute and recovery stages underscores the complexity of the immune response.

### Association between monocyte responses, HPSE expression and EC shedding during acute PUUV-HFRS

To gain further understanding between the relationships of monocyte responses and EGC shedding, we investigated the associations between monocyte frequencies, their HPSE surface expression, ECs (including both CECs and CEPs), soluble plasma HPSE levels and urinary HPSE and syndecan-1 (a heparan sulfate proteoglycan and substrate for HPSE) levels during acute PUUV-HFRS. The data for the latter three parameters was published previously (Cabrera et al., 2022). Our findings revealed a significant association between monocyte distribution and plasma HPSE levels. Decreased frequencies of CMs and increased frequencies of IMs and NCMs associated with increased soluble HPSE levels by spearman correlation analysis, which was corroborated by regression analysis for CM and IM (Fig. 3A-C). Furthermore, increased frequencies of NCMs associated with increased frequencies of ECs, while the NCM surface expression of HPSE associated with less ECs and increased urinary syndecan levels (Fig. 3D-F). While the simultaneous loss of NCMs from the circulation makes the interpretation the observed significant associations rather complex, these findings suggested that NCMs (and potentially IMs) shed HPSE during acute PUUV-HFRS, which resulted in increased soluble HPSE levels, increased numbers of ECs and increased amounts of EGC components in the urine. We also analyzed the potential association between EC, monocyte subpopulations and their HPSE expression with several clinical parameters (Suplementary Fig. 2A) and found that NCM not only correlated with disease severity, as previously suggested (Vangeti et al., 2021), but could also explain 19.81% of the variance in disease severity (R^2^ = 0.1981, p value = 0.0258) (Supplementary Fig. 2B). Moreover, a higher frequency of HPSE-expressing NCMs in circulation was strongly linked to less hypotension (Supplementary Fig. 2C-D). On the other hand, although HPSE-expressing CMs correlated with a few clinical parameters, the deeper analyses of these associations through simple linear regressions were not as significant (Supplementary Fig. 2E-G).

**Figure 3.**
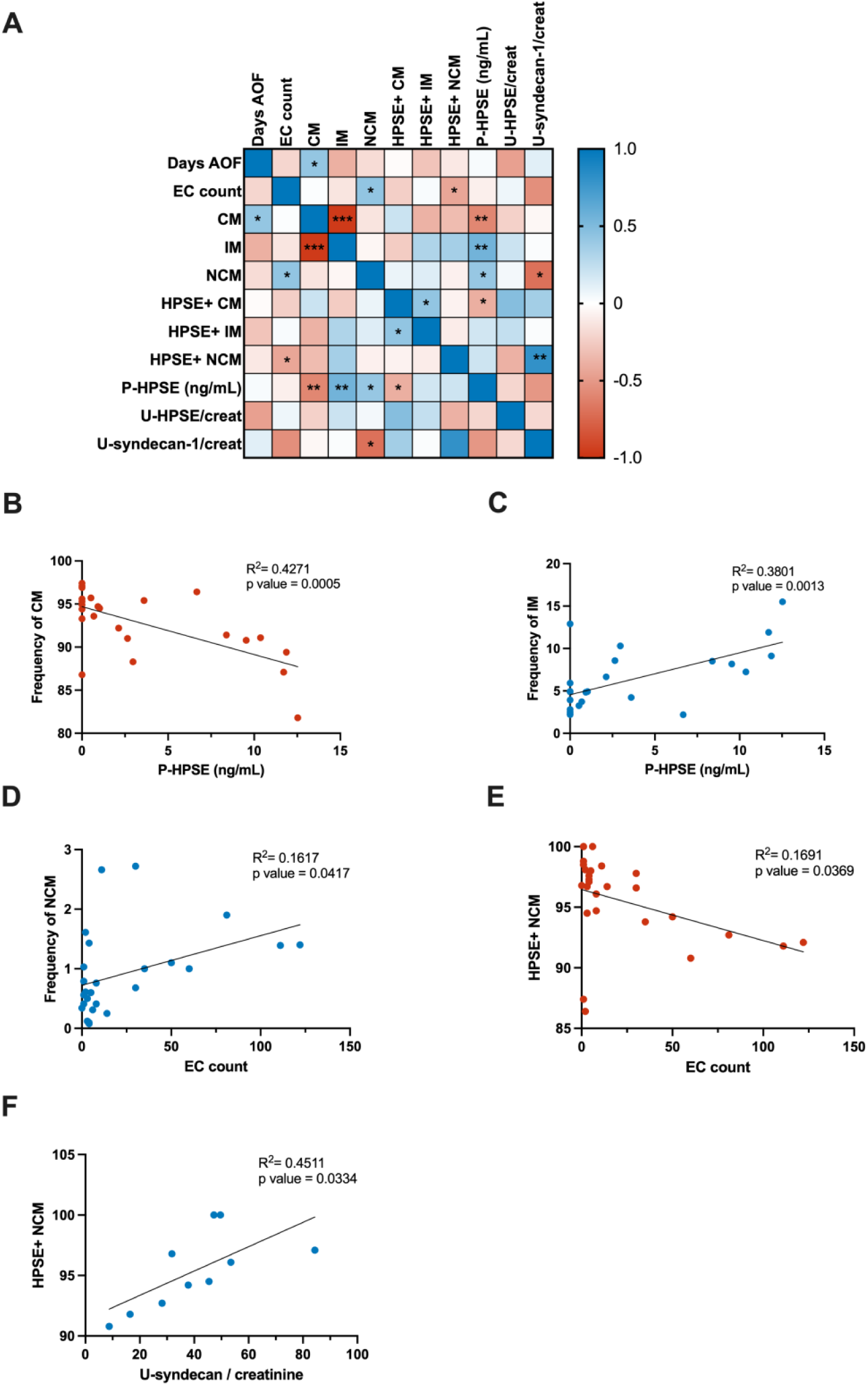
Multiparameter correlation analysis in PUUV-HFRS. (**A**) Correlation matrix depicting the relationships between flow cytometry parameters, including EC counts (including both CECs and CEPs), monocyte subpopulation frequencies, and HPSE expression in monocytes, alongside measured syndecan-1 in urine and HPSE levels in plasma and urine and days AOF. (B) Regression analysis demonstrating a negative association between the frequency of CM and plasma HPSE levels. (C) Regression analysis revealing a positive link between the frequency of IM and plasma HPSE levels. (D) Positive regression analysis displaying the correlation between the frequency of NCM and EC counts from flow cytometry. (E) Regression analysis illustrating the negative correlation between the frequency of HPSE^+^ NCM and EC counts from flow cytometry. (F) Positive regression analysis between HPSE^+^ NCM and urinary syndecan:creatinine ratio. P values were determined using Pearson correlation and are denoted as follows: *p < 0.05, **p < 0.01, ***p < 0.001. *PUUV-HFRS = Puumala orthohantavirus-caused hemorrhagic fever with renal syndrome; HPSE = heparanase; CM = Classical monocytes; NCM = Nonclassical monocytes; EC = endothelial cells; HPSE^+^ NCM = heparanase-expressing nonclassical monocytes; U-syndecan/creatinine = urinary syndecan:creatinine ratio; AOF = after onset of fever*.

### Downregulation of monocyte surface HPSE expression is mediated by PUUV-infected ECs

The observed modulation of HPSE expression in monocyte subsets during acute PUUV-HFRS led us to investigate the direct effect of virus infection in monocyte HPSE expression. Since PUUV is known to productively target ECs, we developed an assay in which isolated human primary monocytes were co-cultured with infected primary ECs before analyzing the surface expression of HPSE in different monocyte subsets as well as ECs by flow cytometry. Approximately 10% of ECs infected with live PUUV but none by UV-inactivated PUUV expressed PUUV nucleocapsid protein which confirmed the presence of virus in the co-culture (Supplementary Fig. 3), prior to the addition of monocytes at 3 days post infection. Interestingly, the surface expression of HPSE significantly diminished in CMs, IMs and NCMs in infected co-cultures as compared to non-infected controls inoculated with UV-inactivated virus (Fig. 4). This finding suggests that virus infection, either directly or indirectly, down-regulated HPSE expression in monocytes. We observed a slight but non-significant downregulation of HPSE expression also in infected ECs.

**Figure 4.**
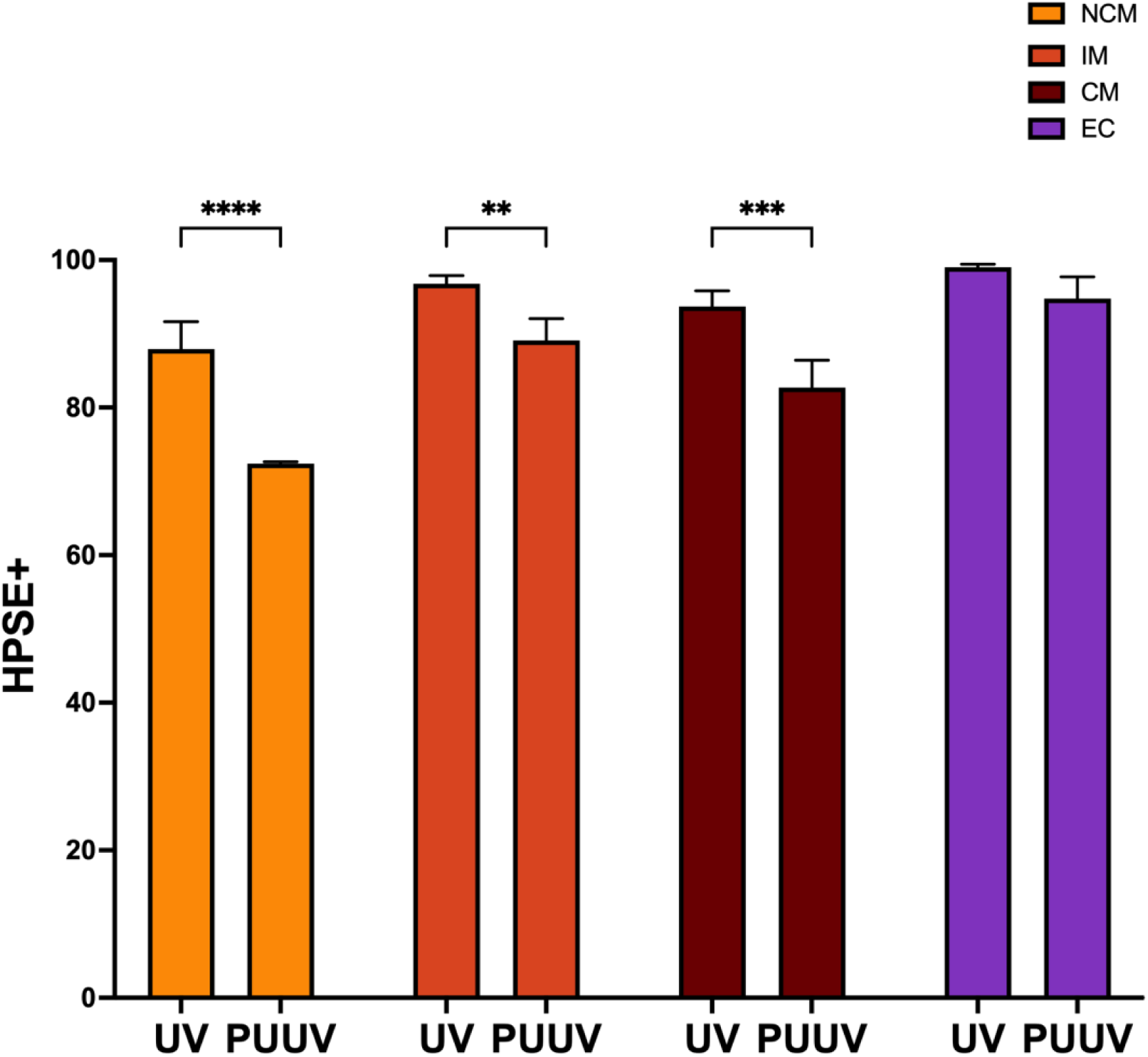
Heparanase expression is decreased in monocyte subpopulations by PUUV-infected endothelial cells. BECs were infected live or UV-inactivated PUUV for 3 days after which monocytes isolated from healthy controls were introduced in the co-culture for 24-hr. Cells were stained with an antibody panel allowing the identification of surface HPSE expression in CMs, IMs, NCMs and ECs by flow cytometry. The statistical differences between UV- and live PUUV-infected co-cultures were assessed by Kruskal-Wallis test. *p < 0.05, **p < 0.01, ***p < 0.001. *PUUV = Puumala orthohantavirus; HPSE+ = heparanase-expressing cell; BECs = blood microvascular endothelial cells*.

### Unsupervised clustering of monocyte populations in acute PUUV-HFRS

Finally, we wanted to gain more insight into the distinct monocyte subpopulations during PUUV-HFRS and employed unsupervised clustering of marker expression, including HPSE, CD138, HLA-DR as well as the well-described CD16 and CD14. By these means we identified 18 distinct monocyte populations during PUUV-HFRS (Figure 5A). Clusters 1, 2, 6, 11, 14, and 18 presented co-expression patterns indicative of IMs, since these clusters showed increased CD16 expression while maintaining significant CD14 expression. These IM clusters displayed variable HLA-DR expressions, suggesting a dynamic transition from CMs to IMs. On the other hand, clusters 3, 4, 5, 7, 8, 9, 10, 12, 13, 15 and 16 exhibited a co-expression pattern marked by higher CD14 levels, and a lack of CD16 expression, thus indicative of CMs. Finally, cluster 17 stands out with elevated levels of CD16 together with a low CD14 marker expression, suggesting a NCM phenotype.

**Figure 5.**
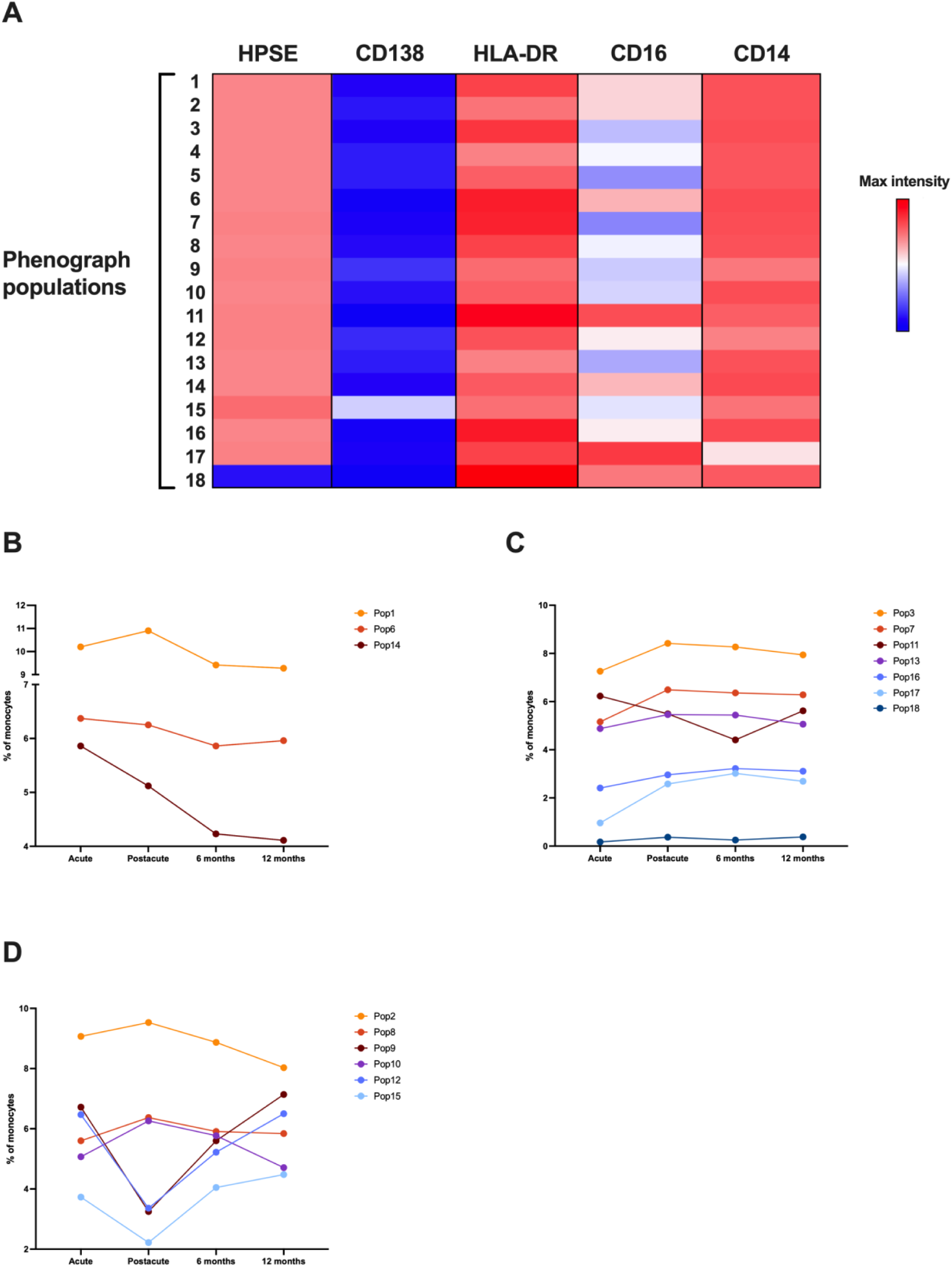
Unsupervised identification of monocyte subpopulations in PUUV-HFRS disease stages. (**A**) A heatmap presenting the MFI of surface markers (HPSE, CD138, HLA-DR, CD16, and CD14) across 18 monocyte subpopulations identified through phenograph clustering analysis. Each row corresponds to a subpopulation, with color intensity indicating the MFI levels of the respective surface markers. (**B–D**) Graphical representation showing the percentage of monocyte subpopulations during different disease phases (acute, postacute, 6 months, and 12 months). Dots connected by line graphs indicate trends in subpopulation changes. (B) Changes in acute and postacute phases are seen in populations 1, 6, and 14 frequencies, which are increased during these phases, compared to recovery. (C) Changes during the acute phase of the disease are detected in populations 3, 7, 11, 13, 16, 17 and 18, where the majority exhibit a decrease in frequency during this phase, while the frequency of population 11 shows an increase. (D) Graph displaying the percentage variations of monocyte subpopulations (pops 2, 8, 9, 10, 12, and 15) specific to the postacute phase in comparison to other phases. *MFI= mean fluorescence intensity; HPSE = heparanase; pop = population; PUUV-HFRS = Puumala orthohantavirus-caused hemorrhagic fever with renal syndrome*.

When analyzing the trends in population changes during different disease stages, we discovered that populations 1, 6, and 14, identified as IMs, showed increased frequencies during both acute and postacute phases (Figure 5B), while other IM populations were either increased during the acute phase (population 11, Figure 5C), or during the postacute phase (populations 2, 8, and 10, Figure 5D). Meanwhile, populations 3, 7, 13, 16, 17, and 18, identified as CMs and NCMs, exhibited decreased frequencies during acute illness (Figure 5C), possibly reflecting their redistribution, as previously described for NCMs (Vangeti et al., 2021), and indicating their potential involvement in response to viral infection. In contrast, populations 9, 12, and 15, identified as CMs, decreased during the postacute phase (Figure 5D), indicating potential shifts in monocyte subsets as the disease evolved.

## Discussion

Investigation of the role of ECs in PUUV-HFRS is critically important, considering their central involvement in vascular responses. ECs play a pivotal role in maintaining vascular integrity, and their dynamic alterations during viral infections can offer crucial insights into the pathophysiological processes. This study aimed to unravel the complex interplay between PUUV infection, innate immune responses, and vascular dynamics by exploring the presence of CECs as marker of EGC degradation and the potential effect monocytes may have on EGC degradation in the circulation of patients during PUUV-HFRS. Our analysis unveiled an evident increase in EC counts and diminished HPSE expression in monocytes during the acute phase of PUUV-HFRS, which gradually returned to baseline levels towards the postacute and recovery stages.

By showing increased levels of CECs and CEPs in acute PUUV-HFRS, our study indicates that EC integrity is compromised during acute disease, as has been speculated by many previous reports (reviewed in (Vaheri et al., 2023)). To our knowledge, this is the first report indicating increased CECs in orthohantavirus-caused disease but is in line with previous reports showing increased levels of circulating EGC components such as syndecan-1 and increased HPSE activity in urine of acute PUUV-HFRS (Connolly-Andersen et al., 2014; Cabrera et al., 2022). Moreover, the persistence of slightly elevated EC counts in postacute PUUV-HFRS suggests ongoing vascular involvement and points towards delayed resolution of vascular damage. The subsequent declining trend in EC counts during the recovery phases may signify a resolution of acute endothelial damage, possibly through the restoration of endothelial integrity or clearance of damaged cells.

A previous study indicated a significant increase in CEPs during the late acute PUUV-HFRS, which is associated with disease severity (Krautkrämer et al., 2014). In contrast to our study, in which we observed the peak of CEP response in the early acute phase (5 days post onset of clinical symptoms), the peak in the CEP response was observed at late acute stage (11-13 days post onset of clinical symptoms). This apparent discrepancy may be due to the differences in the cellular markers used to identify CEPs. Specifically, in our study we classified CEPs as being negative for CD146 despite a previous report indicates that CD146 can be expressed in a subset of CEPs (Delorme et al., 2005). Therefore, our data may underestimate the actual CEP counts and at least partially explain these discrepant data.

Monocytes and their diverse subpopulations play a pivotal role in immune surveillance and response. To explore the origin of CECs, we inspected monocyte subpopulations and their HPSE expression during PUUV infection and recovery. Employing a comprehensive gating strategy, we identified dynamic changes in CM, IM and NCM monocyte frequencies across different stages of PUUV-HFRS, highlighting the complex nature of monocyte responses during viral infections. Particularly, the observed decrease in NCM frequency during acute PUUV-HFRS, which were strongly associated with disease severity, corroborates our previous findings (Vangeti et al., 2021) where we documented NCM redistribution from circulation to affected organs, such as the kidneys, and prompting consideration of the potential local effects of these cells. Interestingly, previous studies on viral diseases have revealed a similar decrease of NCM frequencies during severe COVID-19 (Silvin et al., 2020).

Our further analysis revealed diminished frequencies of HPSE^+^ NCMs in acute PUUV-HFRS. Furthermore, correlation and regression analyses, particularly those involving NCM frequencies and their HPSE expression, revealed a positive association with plasma HPSE activity and negative association with ECs, respectively, implying a potential role of these cells in modulating endothelial responses. This was further supported by the strong power of HPSE-expressing NCMs in influencing blood pressure. These findings are in line with the idea that once NCMs encounter PUUV-infected ECs, their surface expressed HPSE is released to degrade the EGC. Consequently, this degradation could manifest as increased frequencies of CECs and increased EC permeability.

The decreased monocyte HPSE expression by PUUV infection was further corroborated by *in vitro* co-culture experiments involving PUUV-infected ECs and monocytes. These findings demonstrate that the downregulation of monocyte HPSE during HFRS is likely mediated by the virus. Our data does not, however, pinpoint the exact mechanism of HPSE downregulation, which based on the current results could be caused by monocytes directly encountering the virus or by indirect mechanisms mediated by infection-activated ECs. The latter seems more likely due to our previous studies, which indicate increased leukocyte (monocytes and neutrophils) adhesive ability of PUUV-infected ECs as compared to non-infected ECs (Strandin et al., 2018; Vangeti et al., 2021). One argument against the role of virus particles per se in HPSE downregulation is also the strong downregulation of PUUV infection in the *in vitro* infected ECs at the 3-day post infection time point, when monocytes were added to the culture. Only 10 % of ECs showed antigen positivity at this time point, which is significantly less than the initial level of PUUV infection at 1-day time point, as shown previously (Strandin et al., 2016). The decrease in PUUV infection is due to cellular antiviral responses, which could also affect monocyte HPSE expression in co-culture assay setup.

Unsupervised analysis based on surface marker expression elucidated distinct coexpression patterns indicative of CMs, IMs, and NCMs. Monocyte populations exhibiting increased CD16 expression and variable HLA-DR levels were identified as IMs, and their increased frequencies mostly prevailed during both acute and postacute phases, suggesting potential early-stage involvement. This is in line with our previous study, in which we observed strongly increased HLA-DR expression in total IM cell subset in acute PUUV-HFRS (Vangeti et al., 2021). However, it is important to note that this was specific to certain IM clusters identified through phenograph analysis, but not all, since frequencies of certain IM clusters were increased during the postacute phase. It’s crucial to consider these dynamics in the context of the identified monocyte subsets and their distinct roles during PUUV-HFRS progression.

While the overall frequencies of CMs demonstrate relative stability across different disease stages, our detailed analysis reveals population-specific dynamics within CM subsets. Notably, populations 9, 12, and 15, identified as CMs, show decreased frequencies during the postacute phase, suggesting potential shifts in these specific CM subsets as the disease progresses. This nuanced observation highlights the need for a more granular understanding of the distinct responses within CM populations to comprehensively unravel their roles in PUUV-HFRS pathogenesis. Despite valuable insights, a limitation of this study is the lack of tissue-specific analyses, which would offer a more comprehensive understanding.

In conclusion, our study contributes to a better understanding of PUUV-HFRS immunopathogenesis by unraveling EC and monocyte subpopulation dynamics. Observed changes underscore multi-faceted host responses during viral infections, paving the way for research on vascular complications and potential therapeutic interventions. The comprehensive investigation into EC and monocyte dynamics during PUUV-HFRS provides insights into the complex interplay between viral infections, immune responses, and vascular alterations. This investigation prompts additional exploration of the distinct roles played by monocyte subsets in endothelial responses, offering opportunities for interventions in vascular complications linked to PUUV-HFRS. Delving into the contributions of monocytes to endothelial glycocalyx shredding during PUUV infection carries significant clinical implications, as it unveils mechanisms that could be contributing to both vascular leakage and renal damage in HFRS. Therefore, these insights may lead to the identification of novel targets for addressing these complex clinical challenges.

## Data Availability

All data produced in the present study are available upon reasonable request to the authors

## Acknowledgements

This work was financed by grants by the Academy of Finland to T.S. (321809); grants by the Helsinki University Hospital funds to O.V. (TYH 2021343); EU Horizon 2020 programme VEO (874735) to O.V.; Paulon Säätiö to L.E.C.; Suomen Lääketieteen Säätiö to L.E.C.; Finnish Kidney Foundation to L.E.C. and J.T.; Jane and Aatos Erkko foundation to O.V.; and Finnish Cultural Foundation Pirkanmaa Regional fund to J.T. The funders had no role in study design, data collection and analysis, nor decision to publish, or preparation of the manuscript. The authors also thank S. Mäki and M. Utriainen for expert technical assistance.

## Supplementary figure legends

**Supplementary Figure 1.**
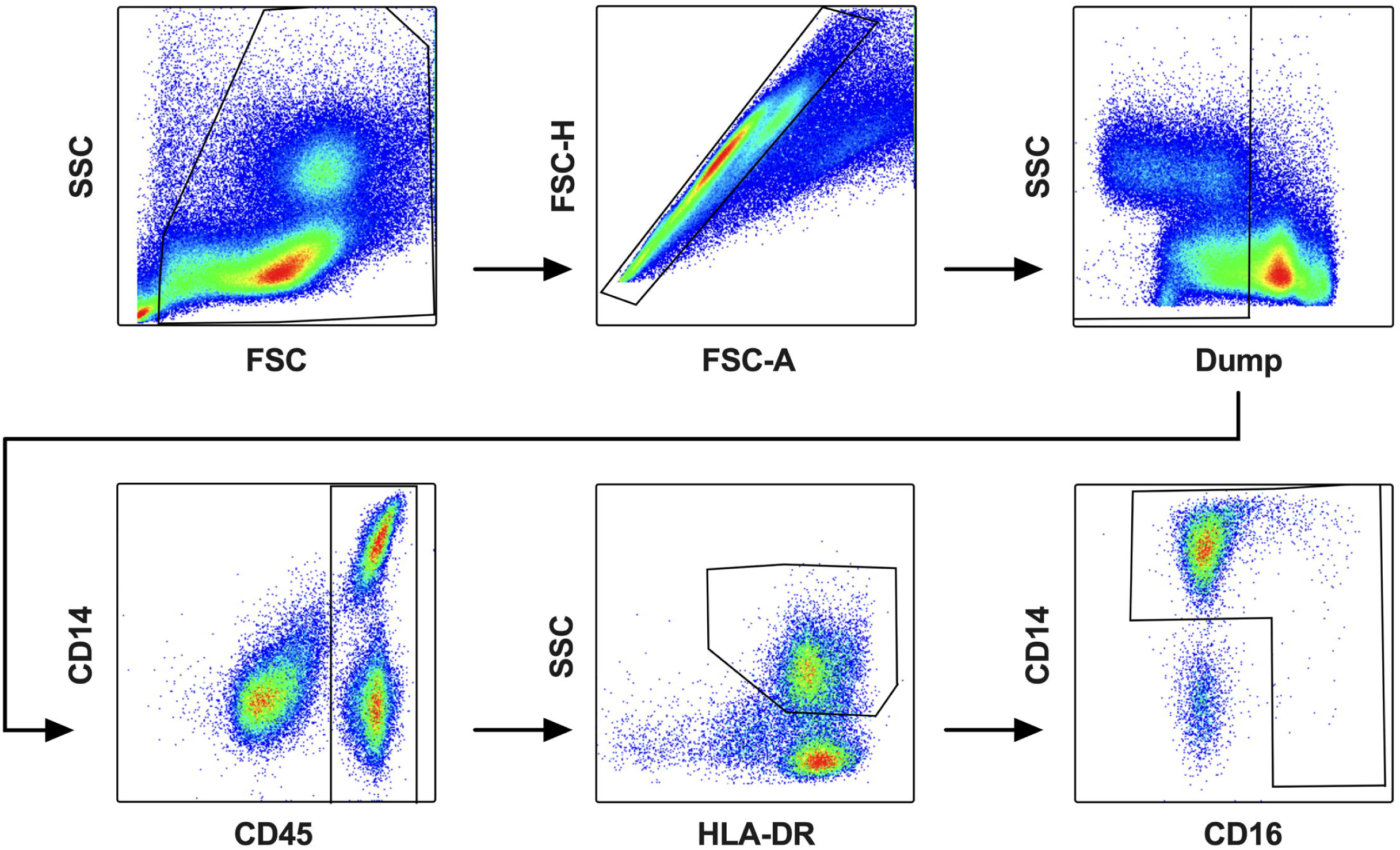
Flow cytometry gating strategy for monocyte identification. Flow cytometry gating strategy outlining the stepwise selection process for identifying monocytes from PBMCs. The strategy consists of six sequential gates to ensure accurate and specific identification: (a) Morphological gating (FSC vs. SSC plot) to exclude debris. (b) Singlet gating to exclude doublets, by plotting FSC area vs. height. (c) Dump channel gate to exclude dead cells and CD3, CD56, and CD19 expressing cell populations. (d) CD45 gating, where the CD45^+^ cell population is selected, represented by CD45 vs. side scatter (SSC) plot. (e) HLA-DR gating, isolating HLA-DR^+^ cells with high side scatter (SSC) to capture monocytes within PBMCs. (f) CD14 vs. CD16 gating, excluding double-negative cells in the CD14 vs. CD16 plot to specifically identify monocytes. *FSC = forward scatter; SSC = side scatter; PBMC =* peripheral blood mononuclear cell; FSC-A = *forward scatter area; FSC-H = forward scatter height*.

**Supplementary Figure 2.**
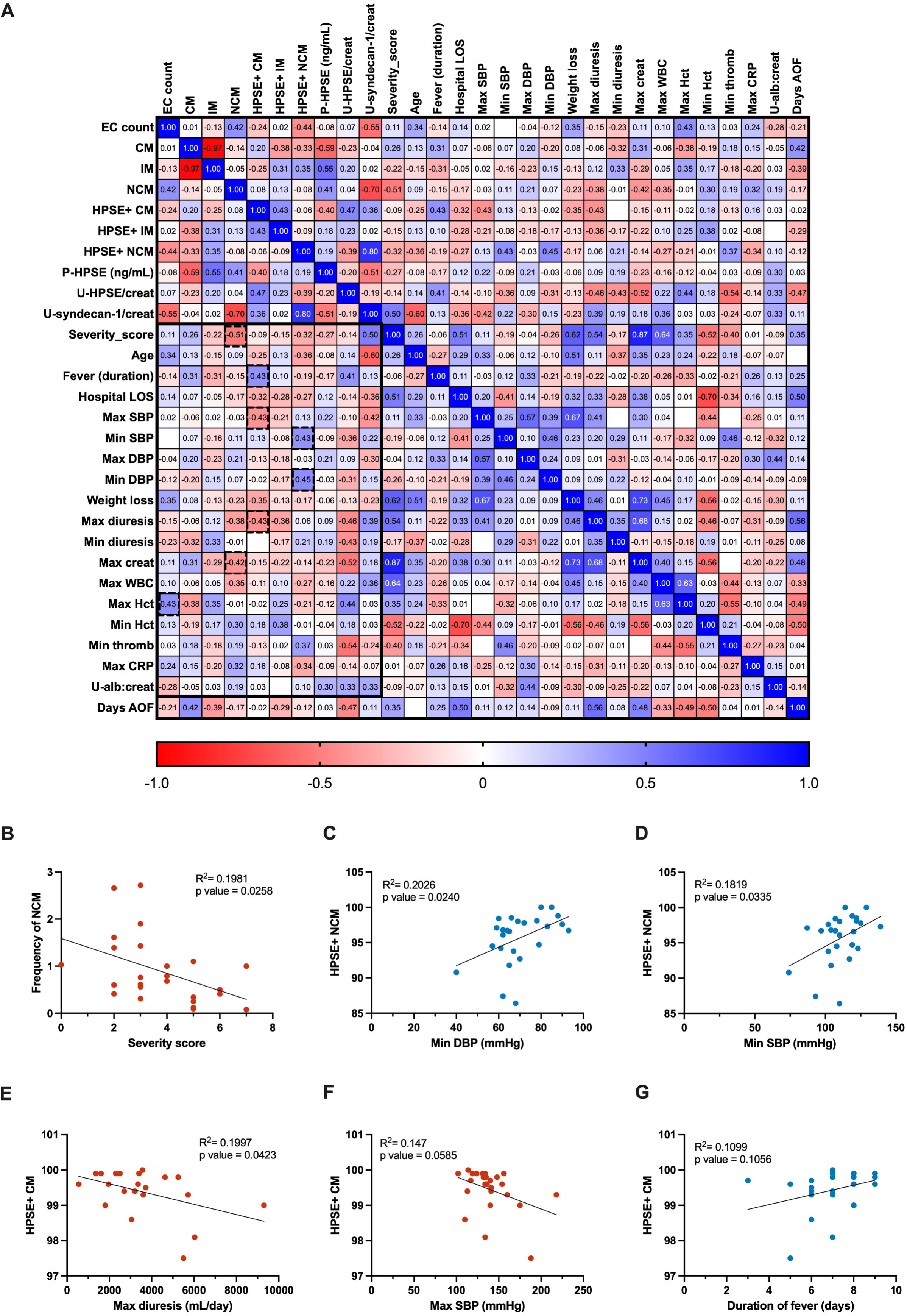
Correlation and regression analyses of ECs, monocyte subpopulations, HPSE expression, and clinical parameters in PUUV-HFRS. (A) Extended correlation matrix depicting the Spearman’s rank correlation coefficients between flow cytometry-derived monocyte subpopulation counts, their respective HPSE expressions, plasma P-HPSE, urine U-HPSE:creat, U-syndecan-1:creat, and various clinical parameters. Clinical parameters are located together and delimited by a black box, where significant correlations (p<0.05) are marked with a surrounding dashed line. (B-G) Simple linear regression analyses illustrating the relationships between: (B) NCM frequency and disease severity score, (C) HPSE^+^ NCM frequency and minimum diastolic blood pressure, (D) HPSE^+^ NCM frequency and minimum systolic blood pressure, (E) HPSE^+^ CM frequency and maximum diuresis per day, (F) HPSE^+^ CM frequency and maximum systolic blood pressure, and (G) HPSE^+^ CM frequency and duration of fever. *EC = endothelial cells; CM = classical monocytes; IM = intermediate monocytes; NCM = non-classical monocytes; HPSE = heparanase; HPSE^+^ = HPSE-expressing cells; P-HPSE = plasma heparanase; U-HPSE/creat = urine heparanase:creatinine ratio; U-syndecan-1/creat = urine syndecan-1:creatinine ratio; LOS = length of stay; min = minimum; max = maximum; SBP = systolic blood pressure; DBP = diastolic blood pressure; creat = creatinine; WBC = white blood cells; thromb = thrombocytes; CRP = C-reactive protein; U-alb:creat = urine albumin:creatinine ratio; days AOF = days after onset of fever*.

**Supplementary Figure 3.**
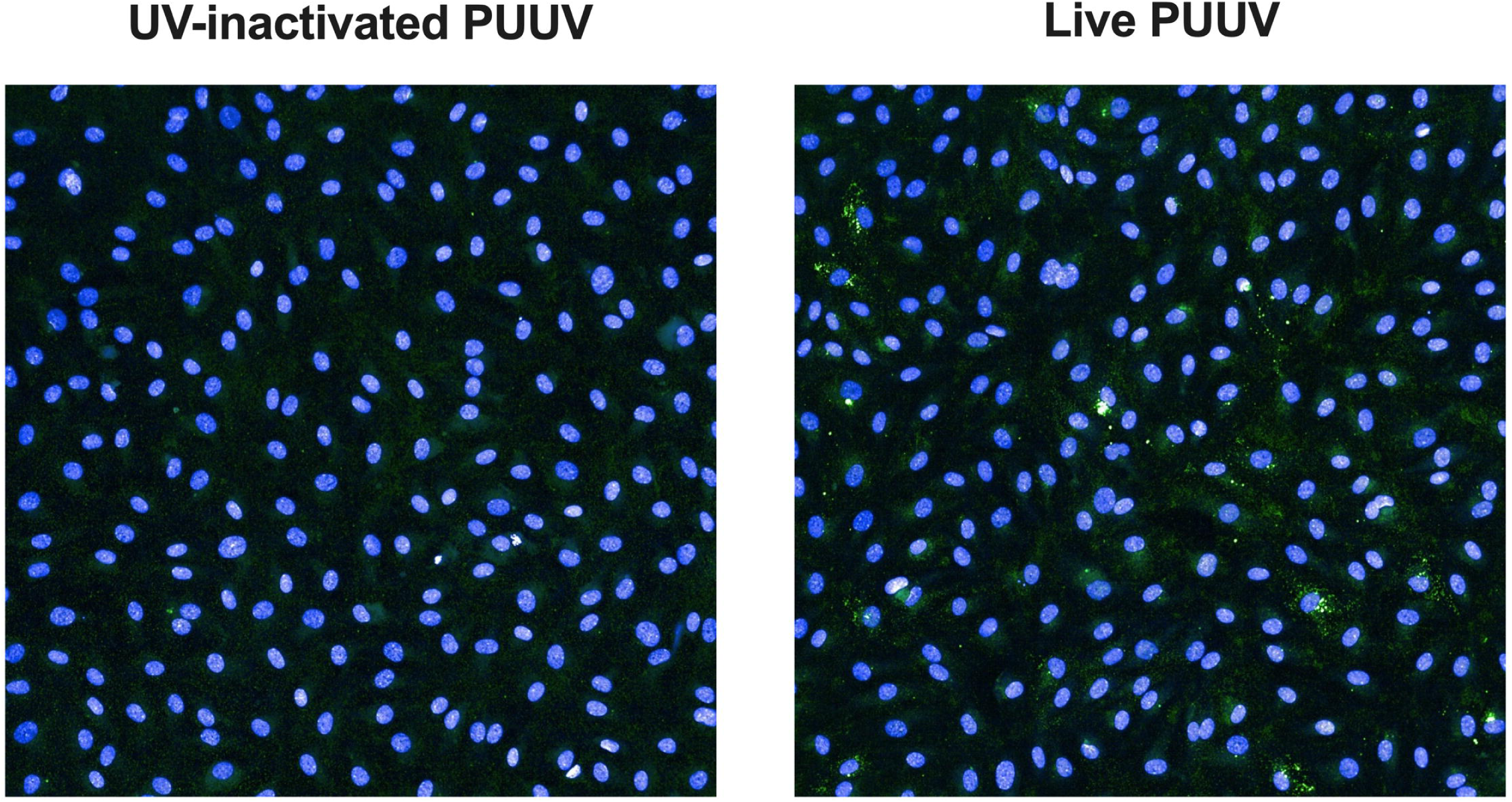
Immunofluorescence of UV-inactivated PUUV and live PUUV-infected blood microvascular endothelial cells (BECs) prior to coculture with monocytes. BECs were infected with live or UV-inactivated PUUV for 3 days and stained for PUUV nucleocapsid protein N and nucleus by PUUV N-specific rabbit sera (green) and Hoechst33342 (blue), respectively. Overlay images of green and blue fluorescence are shown. *UV = ultraviolet; PUUV = Puumala orthohantavirus; N = nucleocapsid; BEC = blood microvascular endothelial cells*.

